# Impaired experience-based attentional suppression in major depression: a neurophysiological and cognitive vulnerability marker

**DOI:** 10.1101/2025.07.28.25332298

**Authors:** Nan Qiu, Peiyang Li, Shaoqing Li, Kena Li, Hongmei Yan, Xianyang Gan, Yanggong Li, Lan Hu, Benjamin Becker, Dezhong Yao

## Abstract

Major Depressive Disorder (MDD) is a leading cause of disability worldwide. Core cognitive deficits, such as impaired selective attention, may contribute to its pathogenesis, yet their neurophysiological basis remains unclear. We here employed an additional-singleton paradigm and examined lateralized ERPs and EEG-based effective connectivity in individuals with MDD (*n* = 33), as compared to healthy controls (HCs; *n* = 31) to examine impaired statistical learning (SL) of distractor locations, a mechanism supporting proactive attention suppression. While HCs demonstrated typical SL effects including faster reaction times and reduced suppression-related Pd amplitudes at high-probability distractor locations, MDD patients failed to benefit behaviorally from SL and instead exhibited enhanced N2pc amplitudes reflecting attention-related selection to high-probability distractor locations, and increased fronto-parietal-occipital connectivity in alpha and theta bands, suggesting maladaptive attentional engagement and compensatory network hyperactivation. Crucially, more negative N2pc amplitudes and earlier latencies were significantly associated with slower cognitive flexibility and greater suicidal symptomology in MDD, independent of medication status. These findings identify a behavioral and neurophysiological candidate mechanism of impaired SL-guided proactive selective attention in MDD. Dysfunctions in SL-guided attentional suppression may represent a neurophysiological phenotype of cognitive vulnerability in depression with potential utility for biomarker and treatment development.

## INTRODUCTION

Major Depressive Disorder (MDD) is a highly prevalent mental disorder and a leading cause of disability worldwide, with prevalence rates continuing to rise globally [1, 2]. Behavioral and neural deficits in emotion- and reward-related processes have been extensively examined [e.g., 3, 4], yet the mechanisms underlying symptomatic and potentially pathogenetic cognitive impairments in individuals with MDD [5, 6] are not well understood. In particular, impairments in basic processes, such as selective attention, may represent alterations that contribute to the development and maintenance of the symptoms [7, 8].

Selective attention enables individuals to prioritize goal-relevant information while suppressing distractors, supporting adaptive behavior in complex environments [9]. Despite its critical role in cognitive and affective processes [10], attentional deficits have received limited attention in MDD, which has traditionally focused on reward-related or affective symptoms. Impaired selective attention may contribute to functional impairments in a wide range of cognitive and affective domains in MDD. Several studies reported that MDD individuals are more easily distracted by task-irrelevant negative stimuli such as sad faces [11–13]. However, attention capture by non-emotional distractors has also been observed, suggesting broader deficits in cognitive control and selective attention [14–16]. For instance, in a non-emotional color-word Stroop task, MDD patients exhibited slower responses, reduced functional connectivity (FC) within the frontoparietal attention network and decreased parietal resting-state alpha power, suggesting an attention-related cognitive vulnerability marker for depression [14]. These findings indicate that attentional impairments in MDD are not limited to emotional biases but reflect generalized attention impairments independent of stimulus type.

Selective attention is traditionally shaped by top-down (goal-driven) and bottom-up (stimulus-driven) mechanisms [17]. Physically salient stimuli can capture attention automatically [18–20], but top-down control enables proactive suppression [21–23]. Beyond this dichotomy, selection history (history-driven) mechanisms has emerged as a third influence [23–26]. SL enables observers to implicitly learn spatial regularities of distractor occurrence, shaping attentional priority [27–30]. In the additional singleton paradigm [31], participants identify a shape-defined target (e.g., a circle among diamonds, or vice versa) while ignoring a color-defined distractor (e.g., a red or green singleton different in color from other items) that appears more frequently at one location [32]. Over time, participants implicitly learn spatial regularities, resulting in reduced interference at the high-probability location (distractor-location effect) and slower responses when targets there (target-location effect) [33–36]. While well established in healthy individuals, such adaptive learning may be impaired in MDD, reflecting deficits in extracting environmental regularities and cognitive control [37–39]. This study investigates whether MDD can implicitly suppress distractors based on spatial SL, offering insights into disrupted attentional learning and informing neurocomputational models and targeted interventions for restoring adaptive, experience-driven attention control.

At the neurophysiological level, two lateralized event-related potential (ERP) components N2pc and Pd reflect attentional deployment and suppression, respectively. The N2pc (∼150– 350 ms), a posterior negativity contralateral to an attended item, indicating selective attention to targets [40–43]. It is sensitive to target saliency and spatial probability, for example, a larger N2pc is typically observed both when targets are more salient (e.g., in pop-out displays) and when they appear at the high-versus low-probability location [44]. The Pd component is a contralateral positivity peaking ∼100–400 ms, reflecting distractor suppression [45]. The Pd was markedly reduced when distractors appeared at the high-probability location, suggesting the reduction of attentional priority of a salient stimulus [46]. Together, N2pc and Pd components offer temporally precise indices of whether attention is captured or proactively inhibited, thus serving as reliable neurophysiological signatures of selective attention, and potential biomarkers of attentional dysfunction in depression.

Beyond ERPs, MDD is associated with disrupted FC, especially in fronto-parietal networks crucial for attentional control [47, 48]. fMRI studies show the importance of the fronto-parietal network in the neural circuitry of depression [49], identifying it as a key region linked to impairments in attentional regulation and executive functioning observed in depressive states [48, 50]. Although fMRI show spatially precise abnormalities, EEG provides superior temporal resolution to track dynamic attentional processes. Theta (4–7 Hz) and alpha (8–13 Hz) oscillations are key to cognitive control and attentional suppression, yet findings on FC alterations in MDD remain mixed [51, 52]. Some EEG studies have reported increased FC in MDD compared to HC in theta oscillations [54], while others have demonstrated the opposite pattern. For instance, Huang et al. [55] observed significantly reduced theta-band coherence in the frontal cortex with MDD. Alpha oscillations are primarily associated with top-down attentional control and the suppression of task-irrelevant information [56]. Fingelkurts and colleagues revealed higher alpha-band FC in MDD [56, 57], while other investigations reported decreased alpha-band connectivity [58, 59]. However, conventional EEG-FC only measures statistical associations and lacks directional information, task-based directional EEG-FC in MDD remains poorly understood.

In this study, we aimed to investigate the neurocognitive mechanisms underlying selective attention deficits in MDD by integrating lateralized ERPs and frequency-specific effective connectivity. To this end, we employed the additional singleton paradigm, participants identified a shape singleton target while ignoring a salient color singleton distractor, which appeared more frequently in one location. Extending previous findings, the present study addresses the following: (1) It incorporated both temporal ERP markers (N2pc and Pd) and spectral Partial Directed Coherence (sPDC) network dynamics, offering a comprehensive assessment of attentional suppression; (2) It explored whether frequency-specific connectivity in the alpha and theta bands, linked to selective attention processes, differentiates MDD patients from healthy controls; (3) It evaluated the potential of EEG-based features to serve as biomarkers of cognitive vulnerability in depression.

## METHODS

### Participants

A total of 40 MDD patients and 31 age- and sex-matched HCs were recruited. Seven MDD patients were excluded due to excessive behavioral error rates (>30%) or substantial EEG artifacts. All participants were right-handed, had normal or corrected-to-normal vision, had normal intellectual functioning, no auditory impairments, and no major medical or psychiatric history. MDD patients were recruited from the Clinical Hospital of Chengdu Brain Science Institute and diagnosed using the criteria in the fifth version of the Diagnostic and Statistical Manual of Mental Disorders (DSM-5) by two experienced psychiatrists, with symptom severity assessed via the 24-item Hamilton Depression Scale (HAMD-24) and Hamilton Anxiety Scale (HAMA), see Fig. 1a. None of the patients met criteria for comorbid anxiety disorders. Medication use was evaluated following standardized procedures, and a total medication load index was computed based on dosage coding (0 = absent, 1 = low, 2 = high), including Escitalopram, Buspirone, and Agomelatine per Physician’s Desk Reference guidelines [59, 60]. The total medication load for each participant was calculated by summing the codes across all medications [59]. Table 1 presents the medication load index for 26 patients; for 7 out of 33 patients medication records were not available. Healthy participants were recruited from the community, had no personal or family history of major psychiatric disorders, and received financial compensation. All procedures were in accordance with the latest version of the Declaration of Helsinki and were approved by the Ethics Committee of the Clinical Hospital of Chengdu Brain Science Institute, University of Electronic Science and Technology of China. All participants provided written informed consent.

**Fig. 1.**
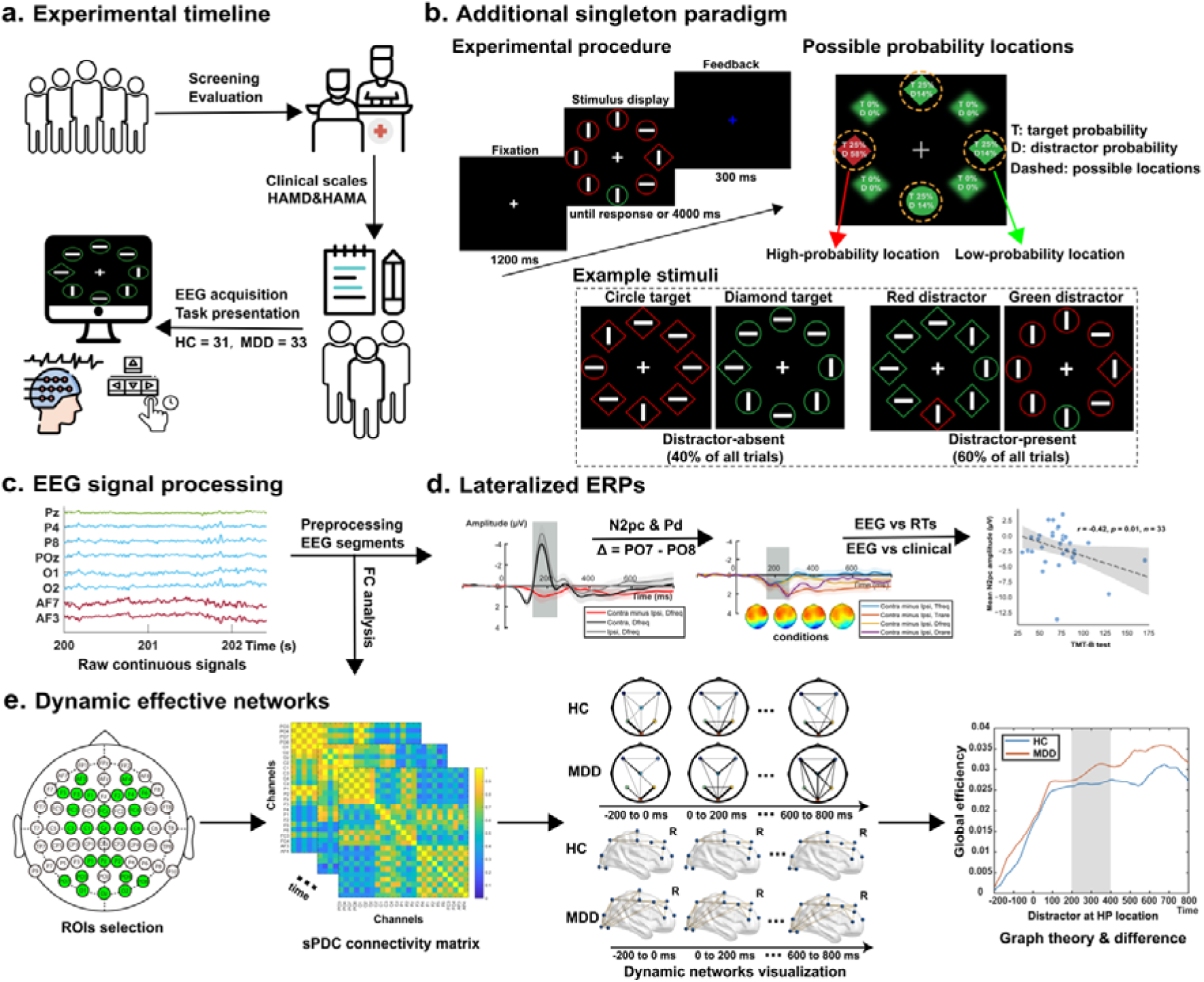
Study design and analytical overview. (a) Illustration of clinical hospital MDD patient recruitment and EEG acquisition. (b) Experimental design. In the additional singleton task, each trial began with a 1200 ms fixation cross and a search display until response or 4000 ms. Participants identified a shape-defined target and reported the orientation (vertical/horizontal) of the line inside it. A feedback display then appeared for 300 ms— white for correct answers, red for errors, and blue for missed trials. For experiment stimuli, in distractor-absent trials, a singleton-shape target—either a circle among diamond-shaped non-targets or a diamond among circle-shaped non-targets. In distractor-present trials, a color-defined distractor was introduced, which could be either a red singleton among green items or a green singleton among red items. For probability locations, the right panel schematically indicates four potential locations (dashed circles not shown during the actual task) where the target and distractor could appear. (c–e) Key EEG analysis steps to (d) isolate N2pc and Pd components and (e) construct time-varying brain networks (see Methods). Images in a–e were obtained from Flaticon.com and Iconscout.com under the free license with attribution.

**Table 1.** Demographic and clinical characteristics.

| Variables (mean ± SD) | HC<br>(*n* = 31) | MDD<br>(*n* = 33) | *t* ($\chi^2$) test | *p* value |
| --- | --- | --- | --- | --- |
| Sex (female\male) | 18\13 | 23\10 | 0.50 | 0.48 |
| Age (years) | 27.71 (±9.87) | 28.09 (±7.92) | 0.17 | 0.86 |
| Education (years) | 15.45 (±1.12) | 15.58 (±0.9) | 1.75 | 0.42 |
| HAMD-24 (score) | – | 24 (±5.95) | – | – |
| HAMA (score) | – | 16.97 (±6.02) | – | – |
| Self-Rating Depression Scale | 40.83 (± 7.25) | 68.59 (± 6.64) | -15.94 | < 0.001 |
| Self-Rating Anxiety Scale | 34.87 (± 5.61) | 59.19 (± 8.85) | -13.21 | < 0.001 |
| Trail Making Test Part A<br>(TMT-A, seconds) | 27.23 (± 7.5) | 29.3 (±11.26) | -0.87 | 0.39 |
| Trail Making Test Part B<br>(TMT-B, seconds) | 56.28 (±13.63) | 76.5 (±33.3) | -3.21 | <0.01 |
| Depressive episodes (number) | – | 1.52 (±0.7) | – | – |
| Duration of illness (months) | – | 38.87 (±33.85) | – | – |
| Duration of first episode | – | 20.31 (±18.56) | – | – |
| Suicide attempts (number) | – | 1 (±0.75) | – | – |
| Suicidal behavior/ideations<br>(number) | – | 2.45 (±0.97) | – | – |
| Self-harm thoughts in the past<br>two weeks (number) | – | 3.12 ($\pm 2.5$) | – | – |
| Self-harm thoughts in the past<br>two weeks (number) | – | 0.5 ($\pm 0.83$) | – | – |
| Antidepressant medications<sup>a</sup><br>(number, %) | – | 14 (42.42%) | – | – |
| Psychotropic medications<sup>b</sup><br>(number, %) | – | 10 (30.30%) | – | – |
| Medication load index<sup>c</sup> (score) | – | 1.1 ($\pm 1.85$) | – | – |

<sup>a</sup> Use of Antidepressant medications: Fluoxetine, Sertraline, Escitalopram, Fluvoxamine, Paroxetine, Venlafaxine, Duloxetine, Vortioxetine.
<sup>b</sup> Use of other Psychotropic medications. Anxiolytic: Tando spirone, Buspirone; mood-stabilising: Sodium valproate sustained-release tablets; Antipsychotic: Quetiapine; Benzodiazepines: Lorazepam, Oxazepam, Alprazolam, Clonazepam; Others: Agomelatine, Trazodone.
<sup>c</sup> The medication load index, based on Sackeim (2001), was calculated for 26 MDD patients by scoring each drug as 0 (absent), 1 (low), or 2 (high); data were unavailable for 7 patients.

### Study design

The experiment was conducted in a dimly lit, sound-attenuated, and electrically shielded room. Stimuli were presented via PsychoPy 2023.2.3 on a 24-inch monitor (1920×1080, 60 Hz). Participants sat 60 cm from the screen and were instructed to remain relaxed to minimize EEG artifacts. Each search display (Fig. 1b) comprised eight colored outline shapes (circles or diamonds) evenly spaced around an imaginary circle (radius: 4°). Circles had a 2° diameter; diamonds measured 2° × 2°. Each shape contained a white line (0.3° × 1.5°, RGB: 255/255/255) oriented vertically or horizontally. On each trial, the target was a shape singleton (one circle among seven diamonds or vice versa), with remaining items serving as non-targets. On distractor-present trials, one non-target was a color singleton distractor (either red [RGB: 255/0/0] among green [RGB: 0/128/0] shapes, or vice versa). Displays were shown on a black background (RGB: 0/0/0) with a central white fixation cross (1° × 1°, RGB: 255/255/255) throughout each trial.

Each trial began with a fixation cross (1200 ms), presented until response or a 4000 ms limit (Fig. 1b). Feedback (300 ms) was given via fixation cross color: white (correct), red (error), or blue (miss). Trials were separated by a jittered interval (0–350 ms). Participants were instructed to fix centrally and indicated the orientation of the line inside (vertical/horizontal) using upward/leftward arrow keys. The task included 1000 trials (10 blocks of 100) and 30 practice trials, lasting ∼40 minutes. In 60% of trials, a distractor was present and in 40% it was absent. The target and distractor could appear at four (top, bottom, left, right) locations. To ensure sufficient lateral distractor-present trials for conducting an EEG experiment, if the target appeared at a midline location, the distractor appeared at the high-probability location with 58% and at each low-probability location with 14%. If the target appeared laterally, the distractor was equally likely among the remaining three. On distractor-absent trials, targets appeared equally across locations. The high-probability location was fixed per participant and counterbalanced. After the task, participants were asked whether the distractors had appeared equally often at all four potential locations or more often at one location. Moreover, they were asked to indicate exactly at which location the distractor had appeared most frequently. In total, fourteen HC and nine MDD participants noticed the uneven distribution, but only one HC and four MDD participants correctly identified the high-probability distractor location, which confirmed SL occurred implicitly and without explicit awareness [37].

### EEG data acquisition and preprocessing

EEG data were recorded from 64 electrodes (10–20 system) using an eego™ mylab system (ANT Neuro), with CPz as reference and GND as ground. Horizontal and vertical EOGs were recorded via F9/F10 and Fp1/inferior orbit electrodes, respectively. Impedances were kept below 5 kΩ. Raw signals were filtered online (0.1–250 Hz), digitized at 2000 Hz, and down-sampled to 500 Hz for further analysis. Data preprocessing in MATLAB (2021a), re-reference to “infinity” zero was performed using the reference electrode standardization technique [61, 62]. Subsequently, data was band-pass filtered by 0.1–30 Hz and cleaned using ocular ICA to remove eye-related artifacts. After continuous EEG preprocessing, segments were extracted from −200 to 800 ms relative to search display onset and baseline-corrected. Trials with incorrect responses or artifacts (e.g., amplitudes > ±60 μV, step > 50 μV/sample, or flatline < 0.5 μV over 500 ms) were excluded.

### ERP Analysis

To examine lateralized ERP components of interest (N2pc, Pd), EEG segments were averaged separately for contralateral and ipsilateral parieto-occipital electrodes (PO7 and PO8) relative to the high-probability location for each condition (see Fig. 2 a-c), and N2pc and Pd were isolated by subtracting ipsilateral ERPs from corresponding contralateral ERPs at PO7 and PO8 electrodes, in accordance with previous studies [63]. Time windows were defined based on both visual inspection of grand-averaged waveforms across groups and conditions, and also following the criteria used in previous studies [64, 65]. Therefore, N2pc was quantified as the mean amplitude within a ±30 ms time window centered on the most negative peak occurring between 150–350 ms post-stimulus. Similarly, the Pd component was quantified using the mean amplitude within a ±30 ms window centered on the most positive peak within the same 150–350 ms time window.

**Fig. 2.**
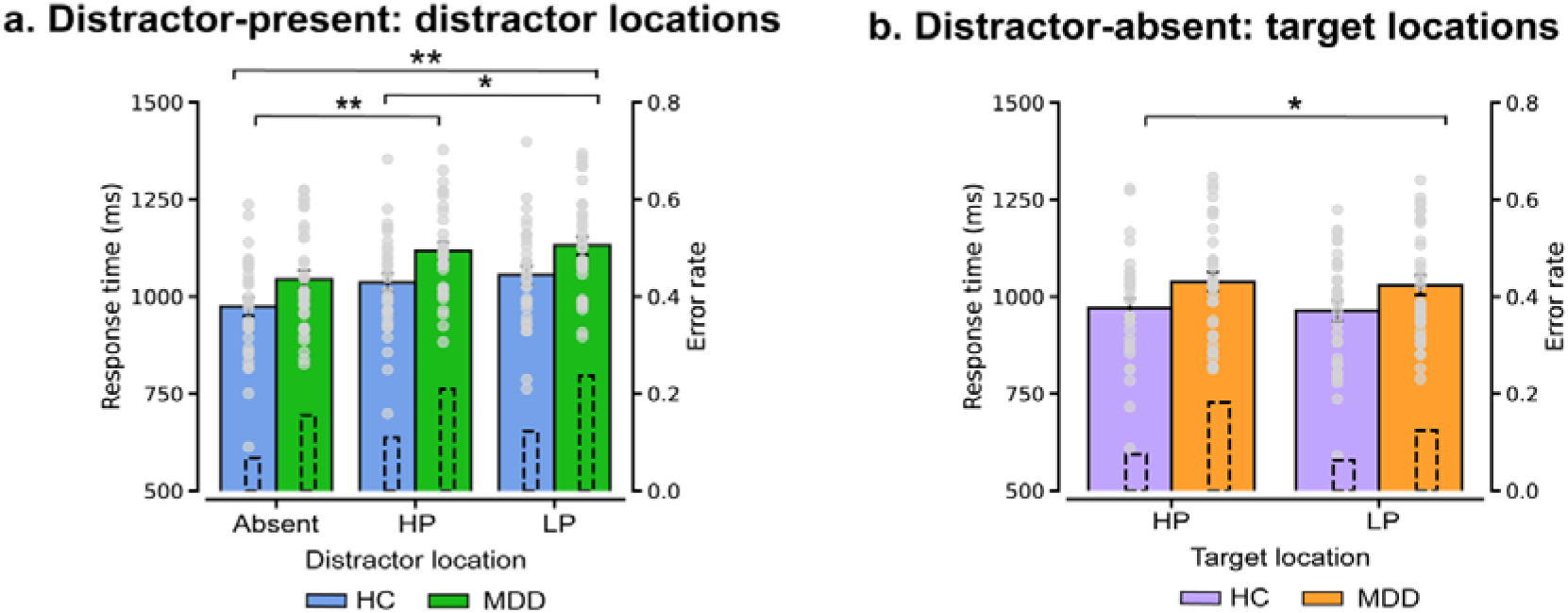
Behavioral results. (a) Mean RTs (solid bars) and error rates (dashed bars) for distractor-present trials, presented separately for three conditions: Absent (distractor-absent), HP (distractor at the high-probability location), and LP (distractor at the low-probability location) for HC and MDD groups. (b) Mean RTs (solid bars) and error rates (dashed bars) for distractor-absent trials, categorized by whether the target appeared at a high- or low-probability distractor location for HC and MDD groups. Error bars represent SEMs, while scatter plots display individual scores, * *p* < 0.05, ** *p* < 0.001.

### Dynamic functional network connectivity analysis

Taking advantage of the sPDC dynamic brain network in analyzing the dynamic evolution of brain information processing [66]. The current study selected 26 electrodes (F1, F2, F3, F4, F5, F6, AF3, AF4, FC3, FC4, Fcz, C1, C2, C3, C4, Cz, P1, P2, Pz, O1, O2, Oz, PO3, PO4, PO7, PO8), covering the whole brain based on the 10–20 system [67] to explore dynamic FC across broad brain regions. Preprocessed EEG segments corresponding to these electrodes were used to construct time-varying connectivity matrices across four task conditions (distractor-present trials: distractor at high- vs. low-probability location, distractor-absent trials: target at high- vs. low-probability location). Connectivity was averaged within theta (4–8 Hz) and alpha (8–12 Hz) bands. On the basis of completing the sPDC estimation of EEG dynamic network, in order to further analyze the dynamic evolution pattern of brain information processing associated with distractor inhibition based on probabilistic cues, dynamic brain network sequence from −200 to 800 ms post-stimulus period was divided into five 200 ms segments. The frequency of occurrence of each edge in each segment is calculated as the strength of the network and normalized. Subsequently, based on the size of the connection strength, the top 10% of the strongest connections are screened and set to 1, and the rest are set to 0. A “zero” value indicates no coupling occurs, and a value of “one” tells perfect phase locking. This binarized dynamic network reflects evolving connectivity patterns during distractor suppression. Finally, global efficiency was computed using the Brain Connectivity Toolbox as the average inverse shortest path length [68], to quantify brain’s capacity for information integration and support group comparisons.

## RESULTS

### Demographics, psychometric and behavior results

Groups were matched for age, sex, and education levels (*p*s > 0.42). The MDD group demonstrated significantly higher symptom load for depression and anxiety (Self-Rating Depression Scale, Self-Rating Anxiety Scale) and impaired executive performance (Trail Making Test Part B, TMT-B; *p*s < 0.01), while basal attention was preserved (Trail Making Test Part A, TMT-A, *p* = 0.39). Trials with extreme response times RTs (< 150 ms, > 4000 ms, or outside the interquartile range), outliers (0.31%) and error trials (12.41%) were excluded from the condition-mean RTs analyses, *t*-tests showed no difference in outlier rate (*t* = −0.83, *p* = 0.41) between groups. MDD patients showed a higher error rate over all trials (*t* = −2.50, *p* < 0.05). See Table 1 for demographic and psychometric details.

#### Distractor interference and SL effects

Repeated-measures ANOVAs examined distractor-location effects on RTs and error rates in HC and MDD groups. As shown in Fig. 2a, RTs were significantly affected by distractor presence in both groups (HC: *F*(2, 60) = 57.28, *p* < 0.001, *η*^2^*_p_* = 0.66; MDD: *F*(2, 64) = 47.45, *p* < 0.001, *η*^2^*_p_* = 0.60). Post-hoc analyses showed RTs were faster in distractor-absent than distractor-present conditions (HC: Δ ≥ 61.82 ms; MDD: Δ ≥ 74 ms, *p*s < 0.001), indicating distractor interference. HCs showed reduced interference at high- vs. low-probability distractor locations (Δ = 18.75 ms, *p* < 0.05), while no such effect was observed in MDDs (*p* = 0.17), suggesting impaired spatial suppression in MDD. Error patterns mirrored RTs, ruling out speed–accuracy trade-offs. On distractor-absent trials (Fig. 2b), MDDs had higher error rates when targets appeared at high-probability locations (13.3% vs. 10.97%, *p* < 0.05), a pattern not observed in HCs (7.45% vs. 6.35%, *p* = 0.30).

#### Group comparisons in SL effects

To further investigate probability cueing effects in MDD, a 2 (Group) × 3 (Distractor Location) repeated-measures ANOVA showed that a significant main effect of Group, *F*(1, 62) = 5.55, *p* < 0.05, η²L = 0.08, and MDDs had slower RTs than HCs (Δ = 76.24 ms, *p* < 0.05). The Main effect of Distractor Location was significant *F*(2, 124) = 100.78, *p* < 0.001, η*²*□ = 0.62). RTs were fastest on distractor-absent trials, slower with high- and lowest with low-probability distractors (Δ_high-absent_ = 67.91 ms, Δ_low-absent_ = 83.98 ms, Δ_low-high_ = 16.06 ms, *p*s < 0.001). Error rates paralleled the RT findings, excluding speed–accuracy trade-off.

### ERP signatures: Deficit attentional suppression of SL-distractors in MDD

#### Distractor-present trials

We examined distractor-elicited Pd in HCs and N2pc in MDDs on distractor-present trials with lateral distractors and midline targets (Fig. 3a–c). In HCs, Pd amplitude was significantly reduced (*t*(30) = 3.11, *p* < 0.05) and latency earlier (*t*(30) = 2.51, *p* < 0.05) for distractors at high- vs. low-probability locations, indicating more efficient suppression (Fig. 3c, e). In contrast, MDDs showed more negative N2pc amplitudes for high-probability distractors (*t*(32) = 2.39, *p* < 0.05) without significant latency change (*t*(32) = 1.67, *p* = 0.10), suggesting impaired temporal adaptation in attentional deployment based on spatial regularities (See Fig. 3d, e).

**Fig. 3.**
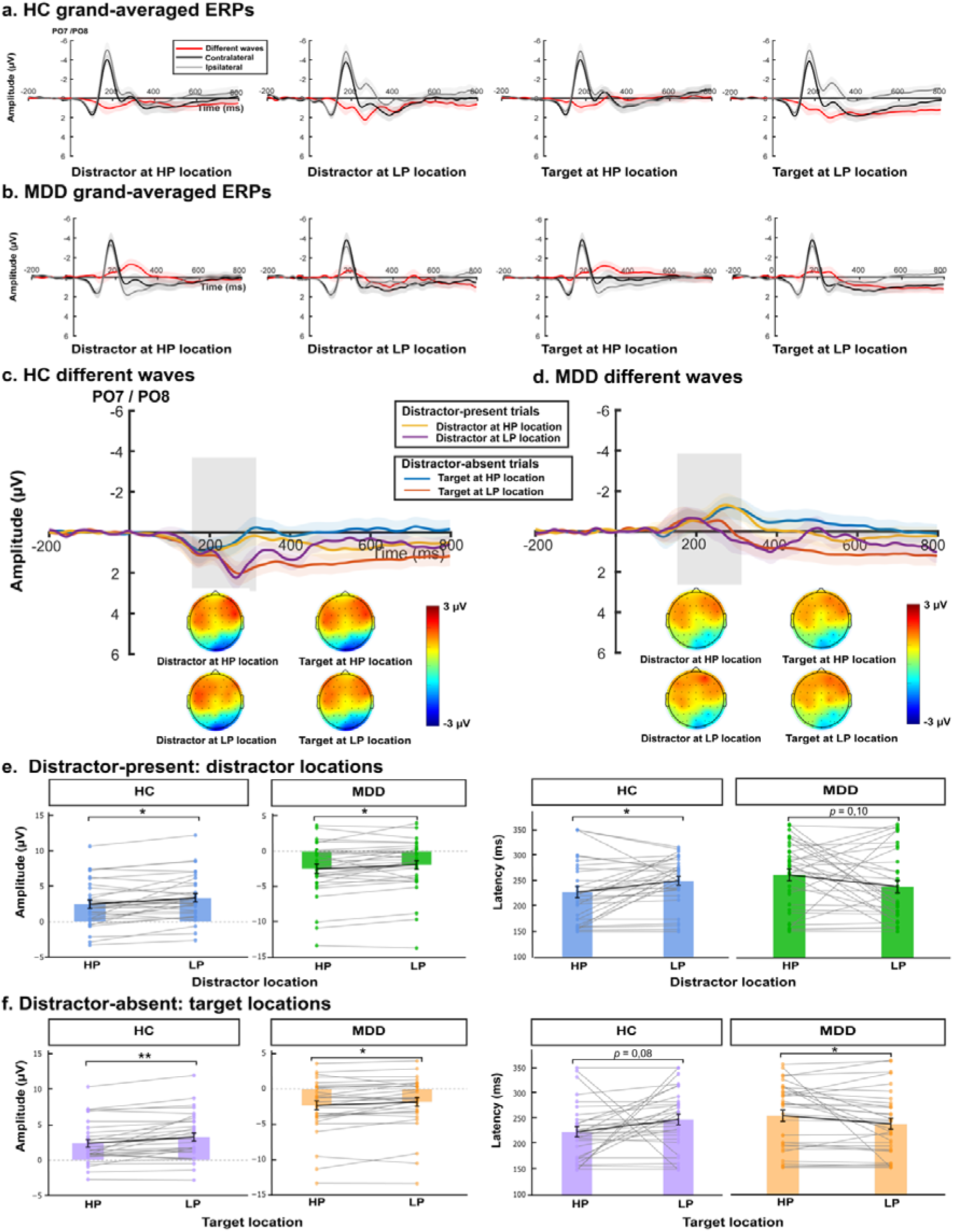
Grand-average ERP waveforms. (a–b) Present distractor- and target-elicited ERPs at electrodes PO7/PO8 as contralateral, ipsilateral, and difference waves from 200 ms pre-stimulus to 800 ms post-stimulus for HP and LP locations in HCs and MDDs, respectively. (c–d) Lateralized difference waves illustrate temporal dynamics of attentional processing under both distractor-present and distractor-absent conditions in HCs and MDDs, respectively. Grey shade marks the 150–350□ms N2pc/Pd window, and topographies show grand-averaged scalp maps (150–350 ms) corresponding to each condition. (e–f) Barplot shows the average amplitudes and latencies of Pd (HC) and N2pc (MDD) for each condition. Error bars indicate SEMs, and scatter plot shows individual value. * *p* < 0.05, ** *p* < 0.001.

#### Distractor-absent trials

In distractor-absent trials, HCs exhibited reduced Pd amplitudes (*t* = 3.60, *p* < 0.001) and marginally earlier latencies (*t* = 1.79, *p* = 0.08) for targets at high- vs. low-probability locations within a 150–350 ms time window (Fig. 3c, f), indicating potential SL-based suppression. In contrast, MDDs exhibited enhanced N2pc amplitudes (*t* = 2.62, *p* < 0.05) and marginally delayed latencies (*t* = 2.02, *p* = 0.05), suggesting persistent attentional allocation to high-probability locations even without distractors, reflecting impaired SL-based suppression.

### Correlation between ERP metrics and clinical symptoms

To explore links between selective attention and depression-related cognitive symptoms, we conducted permutation-based correlations (10,000 iterations) between distractor-elicited N2pc (amplitude, latency) and measures such as medication load, TMT-B performance, and suicidal symptoms in the MDD group. The mean amplitude of distractor-elicited N2pc was not significantly linked with the total medication load index (*r* = −0.05, *p* = 0.79), suggesting that medication effects are unlikely to drive SL-based distractor suppression in MDD (Fig. 4a). However, greater (more negative) N2pc amplitudes and earlier latencies were significantly associated with slower TMT-B performance and more severe suicidal symptoms, suicide attempts and behavior (*r* = −0.42, *p* = 0.01, *r* = −0.37, *p* = 0.03, *r* = −0.33, *p* = 0.06, respectively; Fig. 4b–d). The inefficiency of neuronal N2pc responses may not only affect basal attention processes but also impair higher order executive functions.

**Fig. 4.**
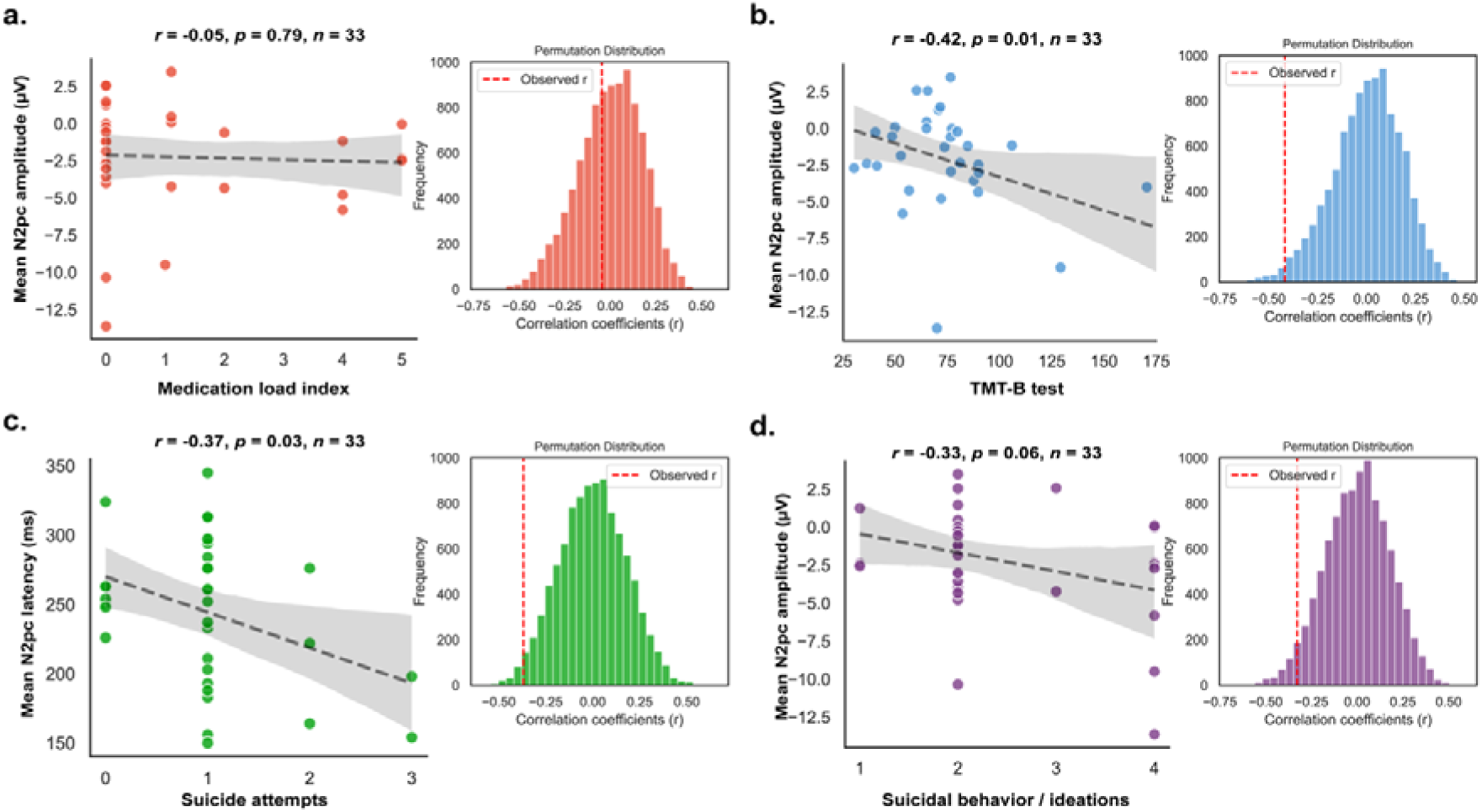
Association between distractor-elicited N2pc and clinical/cognitive symptoms in MDD. (a) MDD patients’ medication load index and N2pc mean amplitudes were uncorrelated. (b) and (c) Scattered plots of negative correlations between TMT-B test and N2pc mean amplitudes and suicide attempts and N2pc latencies. (d) A scatter plot shows a moderately negative connection between N2pc mean amplitudes and suicidal behavior/ideations. Grey denotes the 95% confidence interval. The histograms depict permutation test correlation coefficients, whereas the dashed lines reflect the genuine correlation.

### Dynamic brain networks

We employed the sPDC dynamic network approach to studying brain connection patterns in distractor suppression of SL effects due to its ability to analyse brain information processing evolution. We recorded important information nodes, long-distance linkages, global efficiency, and driving relationships between regions and groups.

#### Group differences in alpha-band sPDC connectivity

We examined group differences in temporal dynamic changes of alpha-band FC across five time windows (−200–800 ms) for distractor-present and distractor-absent trials. In the distractor-location effect (sPDC in distractor at the high-probability location minus distractor at the low-probability location), MDD patients showed increased frontal-to-parietal-occipital and parietal-to-occipital directed FC (Fig. 5a, c). Global efficiency as the critical graph theory analysis indicator (Fig. 5b) was significantly higher in MDD than HC, especially during 200–400 ms (*t*s > 2.63, *p*s < 0.001), suggesting abnormal attention network synchronization. For the target-location effect on distractor-absent trials, MDDs again showed enhanced frontal–parietal–occipital FC and global efficiency in MDDs than HCs from early attention orienting when to the attentional engagement stages (0–400 ms, *t*s > 14.09, *p*s < 0.001), followed by reduced efficiency at 600–800 ms (*t* = 2.19, *p* < 0.05) for targets at high-probability locations, indicating cognitive fatigue. When targets appeared at low-probability locations, MDDs maintained higher efficiency throughout 0–800 ms (*ts* > 3.51.09, *ps* < 0.001), suggesting excessive recruitment of attention resources.

**Fig. 5.**
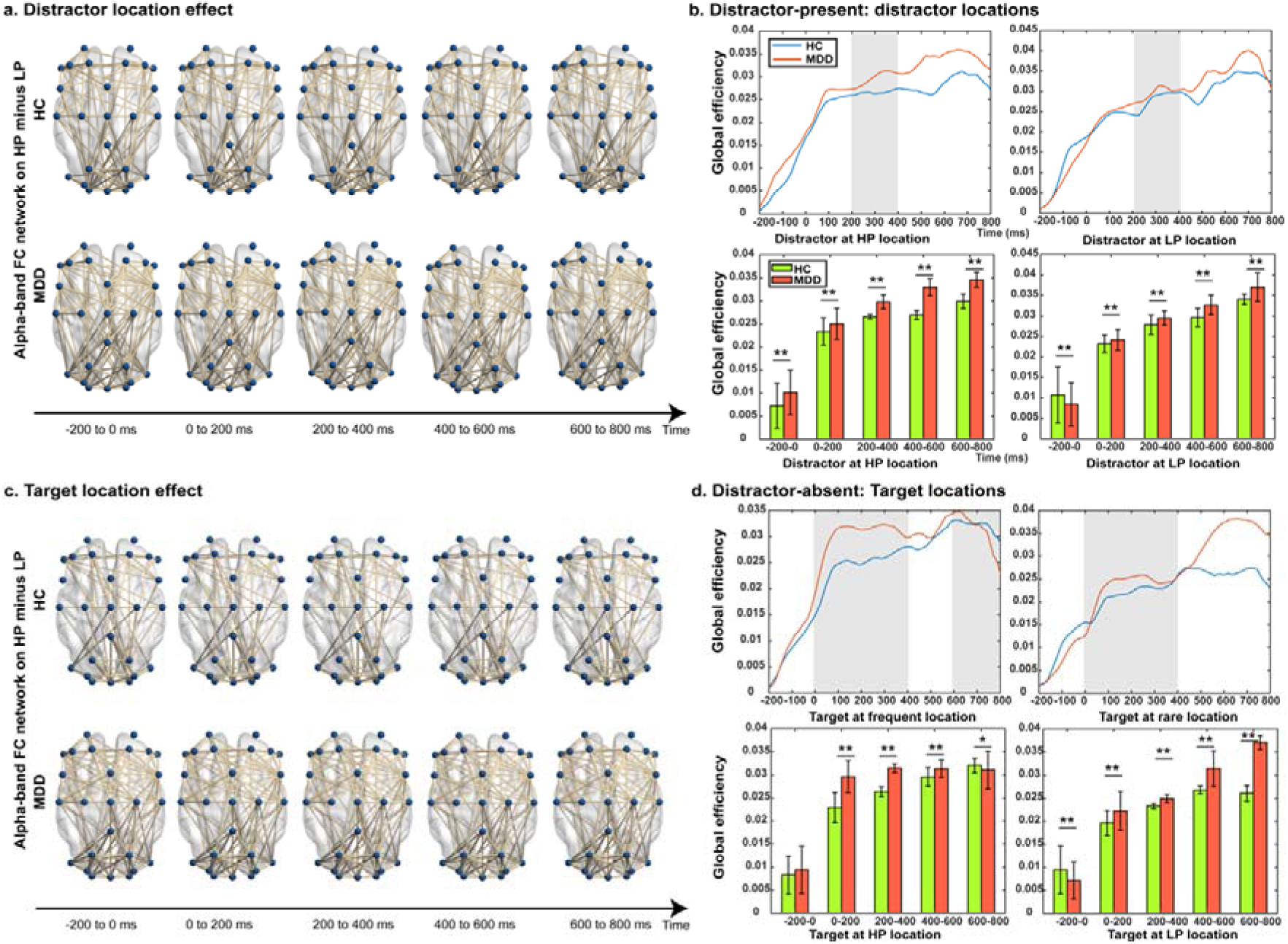
Effective sPDC-based brain network in alpha-band (8–13 Hz). (a) and (c) Comparison of HC and MDD groups for the distractor-location effect (high-probability minus low-probability location) and the target-location effect connections, respectively. The 26-electrode network reveals frontal to parietal and occipital information transfer. (b) and (d) Comparisons of alpha-band global efficiency between HC and MDD groups in distractor-present and distractor-absent conditions for five time-windows, respectively. A two-dimensional waveform plot shows the average alpha power and its grand-average scalp distribution, with grey shades indicating the significant time windows between groups in the upper panels. Bar plot displays the mean global efficiency for each 200 ms time window. Error bar denotes SEMs. * *p* < 0.05, ** *p* < 0.001.

#### Group differences in theta-band sPDC connectivity

In distractor-present conditions (Fig. 6a), MDD patients also showed stronger right frontal-parietal-occipital theta connectivity than HCs across all attentional stages (−200 to 800 ms), suggesting increased cognitive load and hypersensitivity to external stimuli during distractor suppression. For the distractor-location effect, global efficiency (Fig. 6b) was significantly higher in MDDs than HCs for high-probability distractor locations (−200–0 ms, 200–800 ms; *ts* > 3.92, *ps* < 0.001), and low-probability distractor location (0–800 ms, *ts* > 3.04, *ps* < 0.05). Similar to the target-location effect in the alpha-band, MDDs showed increased connectivity and efficiency than HCs (0– 600 ms, *ts* > 3.49, *ps* < 0.001; Fig. 6c-d), especially at 0–400 ms for high-probability and 0– 800 ms when targets at low-probability locations (*ts* > 3.09, *ps* < 0.001). A marked theta-band activity decline at 600-800 ms (*t* = 48.07, *p* < 0.001) when targets at high-probability locations, indicating cognitive fatigue in the later stage.

**Fig. 6.**
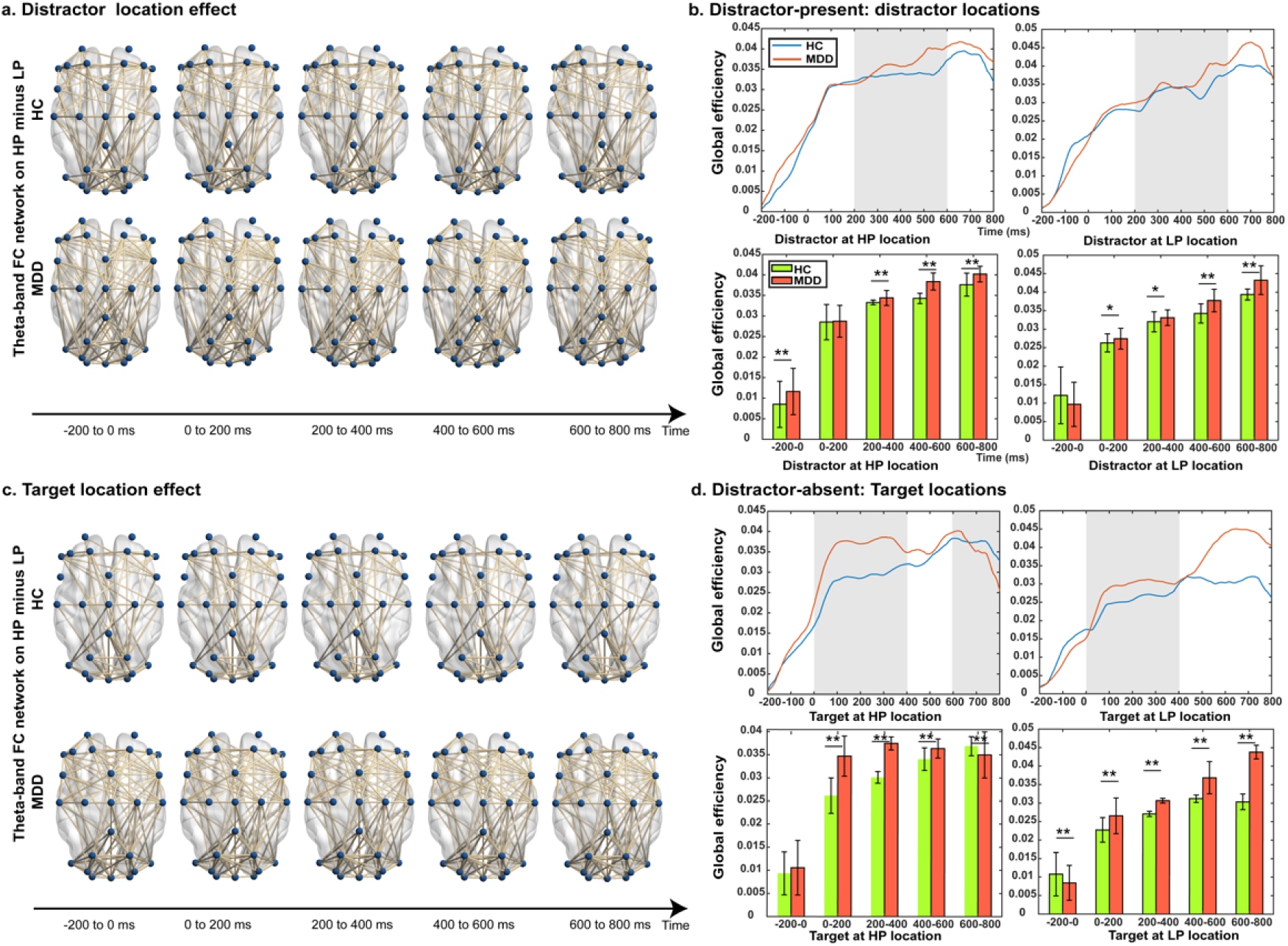
Effective sPDC-based brain network in theta-band (4–8 Hz). (a, c) Show frontal to parietal and occipital information transfer connectivity differences between HC and MDD groups for distractor- and target-location effects, respectively. (b, d) Compare global efficiency between groups under distractor-present and distractor-absent conditions across five time windows. Upper waveform plots depict theta power and shaded regions indicating significant group differences. Lower bar graphs show mean global efficiency per 200 ms window (± SEMs). * *p* < 0.05, ** *p* < 0.001.

Overall, these findings suggest that MDD patients adopt compensatory mechanisms reflected in increased alpha- and theta-band connectivity and global efficiency, notably in fronto– parietal–occipital networks. However, when the target appears in the high-probability location, cognitive compensation ultimatly fails. Due to attentional flexibility and SL impairments in depression, initial overrecruitment and subsequent failure may critially underlie cognitive control deficits. These findings indicate a potential neurophysiological candidate marker for deficits in experience-guided proactive attention in depression.

## DISCUSSION

Using experience-dependent spatial distractor learning we demonstrate specific deficits in proactive selective attention gating in MDD. HC exhibited the typical behavioral and ERP markers of distractor suppression in the additional singleton paradigm combined with EEG: faster RTs and decreased Pd amplitudes for distractors at high- vs. low-probability locations. The MDD group did not show the attention facilitation - RTs did not differ among distractor locations, and distractor-elicited Pd components. Instead, distractors and targets at high-probability locations elicited increased N2pc amplitudes in MDD, suggesting aberrant attentional capture and persistent allocation to these locations even in distractor-absent trials. This reflects impaired proactive suppression in MDD patients. Importantly, ERPs changes were accompanied network-level changes, such that MDD patients exhibited (1) increased sPDC-based directed connectivity in alpha- and theta-band circuits, especially frontal– parietal–occipital, and (2) higher global efficiency in network property evaluations, suggesting neural compensatory recruitment to meet attentional demands. These findings imply that MDD patients over-recruit attention networks but fail to suppress spatial distractors, reflecting impaired neurofunctional utilization of experience-dependent spatial regularities to guide attention.

For HCs, our results align with prior work demonstrating SL reduced distractor interference (reduced RTs) at high-probability locations, reflecting a robust distractor-location probability-cueing effect [28, 33, 34, 69]. Electrophysiologically, SL reduced interference, which was evidenced by distractor-elicited Pd amplitude from salient distractors at high-compared to low-probability distractor locations, consistent with targets elicited Pd on distractor-absent trials, indicating spatial regularity-based proactive suppression. Wang [63] and van Moorselaar [70] also observed target-elicited Pd due to SL-based distractor spatial locations, not feature expectations. Notably, our results support the idea that spatial expectancies are encoded in the Pd component [71, 72]. Accordingly, both distractors and targets at high probability distractor locations elicited an early Pd [63, 73]. Our findings highlight that distractor-elicited and target-elicited Pd reflects a proactive suppression mechanism in the attention priority map with salient stimuli automatically reducing priority signal weight, reducing attentional capture or even proactively supressing it [21, 34, 37].

In contrast, MDD patients failed to exhibit SL-based facilitation effects, but rather exhibited enhanced N2pc amplitudes and prolonged latency to targets at high-probability distractor locations. This may reflecting maladaptive attentional deployment and delayed attentional engagement. Previous studies showed that MDD is associated with impaired selective attention, leading to impaired or biased attention and impaired daily fucntioning [15, 74, 75]. MDDs exhibit distractibility, inability to focus, and inability to multitask [76–78]. Our findings indcate the underlying mechanism by shwoing that the patients exhibt distractor-elicited N2pc across all conditions, while targets at the high-probability location elicited larger N2pc amplitudes. On distractor-absent trials, MDD patients demonstrated delayed N2pc at the high- vs. the low-probability location, suggesting aberrant attention deployment speed. N2pc represents a neurophysiological marker of attentional engagement [79, 80] and of saliency during visual search [81, 41].

We hypothesized MDD exhibits increased attentional allocation at the high-probability distractor location because both distractors and targets are perceived as more salient, indicating maladaptive attentional engagement rather than SL-learned suppression. In the context of our non-emotional distractors, these findings support a general selective attention deficit in MDD [13, 15]. The absence of SL suppression suggests that MDD is associated with reduced sensitivity to selection history and impaired utilization of past experience to guide attention in visual environments. Further, exploratory analyses revealed that greater N2pc amplitudes and earlier latencies were associated with poorer cognitive flexibility and more severe suicidal symptoms in MDD, independent of medication load. This may reflect underlying cognitive dysfunction and serve as a potential neurophysiological marker of generalized attentional deficit and clinical severity in depression.

Effective network analyses is an innovative approach that determines individual differences in the temporal dynamics underlying task-related effective connectivity. Our network-level findings were the first to examine oscillatory-resolved sPDC based on selective attention in MDD patients. If SL-based distractor suppression operates proactively, it should be reflected in brain networks of alpha-band (8–12 Hz) activity, which has been strongly linked to proactive suppression of distractors [34, 63, 72]. In our study, individuals with MDD exhibited abnormal effective connectivity during attentional selection in MDD. Compared to HCs, MDD patients showed significantly increased directed connectivity from the right frontal cortex to parietal and occipital regions across all task-relevant periods, along with sustained fronto-parietal-occipital connectivity from −200 to 800 ms—even in distractor-absent trials—indicating persistent recruitment of attentional control networks. Previous research has shown that increased activation of alternative pathways in the fronto-parietal network in MDD may be compensatory for impaired attention and working memory, requiring greater cognitive control to process information as efficiently as HCs [82, 83]. As reported, global efficiency in the Kinaesthetic Motor Imagery task increased over time for patients with hemiplegic stroke and healthy controls and increased as more advanced cognitive functions in the brain were mobilized [84]. Similarly, theta-band analyses revealed sustained and enhanced right-sided fronto-parietal-occipital connectivity in MDD across the attention period, reflecting increased cognitive load and proactive control. It has been reported that in the theta-band, the global efficiency of the cortical connectivity network increased significantly in the 2-back task compared with the 0-back task during the working memory task, the strengthened theta oscillation and more globally efficient network structure in the theta band might be attributed to the need for enhanced attention to facilitate the sustained maintenance of memory representations [85].

Notably, in the alpha-band sPDC brain networks, for the MDD group, when targets appeared at the high-probability location, global efficiency increased during 0–400 ms, followed by a marked decline during 600–800 ms, suggesting early over-engagement and later cognitive fatigue. In contrast, global efficiency remained high throughout 0–800 ms at low-probability locations in MDD. This may indicate an impaired ability to flexibly modulate cognitive effort based on task context and probabilistic cues. Similarly, in the theta-band, MDD patients showed higher global efficiency during early attention phases (0–400 ms), followed by a sharp decline during 600–800 ms, especially when targets appeared at high-probability distractor locations. According to this drop suggests a collapse of sustained top-down control and the depletion of limited cognitive resources, a pattern that aligns with prior neuroimaging findings of dysfunctional persistence in frontoparietal control systems in MDD [86]. But the current study extends them by revealing the fine-grained temporal dynamics underlying this dysfunction.

Our network-level findings suggest that MDD is characterized by over-engagement and reduced flexibility of attentional networks. Increased connectivity and global efficiency during early stages may reflect compensatory efforts but ultimately lead to inefficiency and cognitive fatigue. Altered alpha- and theta-band dynamics may reflect the cognitive core symptoms of MDD, such as attentional rigidity and difficulty adapting to environmental regularities. These oscillatory connectivity patterns could serve as biomarkers of network-level dysfunction and guide future interventions (e.g., closed-loop brain stimulation). However, EEG’s limited spatial resolution and the scarcity of research on event-related connectivity in MDD constrain current interpretations. Multimodal imaging with larger samples is needed to clarify subcortical contributions to attentional dysfunction in MDD.

Integrating behavioral, ERP, and connectivity approaches, our results reveal that MDD patients exhibit impaired utilization of statistical regularities to suppress irrelevant distractors, reflecting impaired proactive attentional suppression. MDD patients displayed increased N2pc amplitudes at high-probability distractor locations and elevated alpha- and theta-band fronto-parietal-occipital connectivity. The observed alterations in oscillatory connectivity and global efficiency likely reflect compensatory attempts to maintain flexible attentional control. By combining behavioral, electrophysiological, and network-level measures within a statistical learning framework, this study highlights disrupted attentional regulation and inefficient network dynamics as key neurocognitive features of MDD and identifies selective attention impairment as a mechanistically distinct and clinically relevant cognitive phenotype in MDD, which may offer a promising target for circuit-informed, precision interventions aimed at restoring adaptive attentional control.

## Data Availability

The codes and data supporting the findings of this study are available from the corresponding author upon reasonable request and with permission from the university administration.

## ACKNOWLEDGMENTS

This work was supported by STI 2030-Major Projects (No. 2022ZD0208500); STI 2030 Major Projects 2022ZD0211400 (H.C.); National Natural Science Foundation of China (Nos. 82101620); Youth Innovation Project of Sichuan Medical Association (Q23045) and Scientific and Technological Research Program of Chongqing Municipal Education Commission KJQN202200640.

